# Dysphoric Milk Ejection Reflex: Risk, Prevalence, and Persistence

**DOI:** 10.1101/2024.06.25.24309475

**Authors:** Romy Cappenberg, Jesus Garcia Garcia, Christine Happle, Anna Zychlinsky Scharff

## Abstract

**Background:** Dysphoric Milk Ejection Reflex (DMER), which affects a significant proportion of lactating parents and may significantly impact feeding choices, is poorly understood.

**Objective(s):** The primary aim of this study is to characterize prevalence, duration, and factors influencing DMER as well as its effect on parental feeding choices.

**Study Design:** We conducted a cross-sectional study using an online survey of lactating parents of children under the age of 18 months who were nursed for any period of time. N=1469 survey responses were collected, of which n=209 reported experiencing DMER. The DMER sub-cohort was further queried about their specific experiences. We performed a risk factor analysis using logistic regression.

**Results:** The prevalence of DMER in our cohort was 14.2% (n=209/1469). We found a high co-incidence of DMER with both postpartum depression and baby blues. DMER was also associated with a pre-pregnancy mental health history, higher education level, and an immigration background. We found no effect on DMER rates by marital status, household income, BMI, history of medical illness, mode of birth and sex of infant, use of assisted reproductive technology, medication during pregnancy, history of abortion or miscarriage, extended perinatal hospitalization, and spousal parental leave.

Among the n=209 women who reported having experienced DMER, 57.7% (n=113/196) reported their symptoms lasted 1-5 minutes, and the most frequently selected descriptions were: tense, hypersensitive, frustrated, irritable, overwhelmed, sad, lonely, and restless. Most women (n=132/180, 73.3%) reported experiencing DMER only at the beginning of a nursing session, while a minority had DMER at every letdown.

Of the DMER group, 85.9% (n=158/184) used a pump to express breastmilk. Of these 57.0% (n=90/158) experienced milder or absent DMER symptoms while pumping as compared to nursing. Only 5.7%, (n=9/158) reported more severe symptoms while pumping, and 12.0% (n=19/158) experienced symptoms only when pumping.

40.2% (n=72/179), of respondents with DMER reported that their symptoms remained stable between birth and weaning. In 29.6% (n=53/179), symptoms became milder and in 9.5% (n=17/179) they disappeared completely. In contrast, 17.9% (n=32/179) reported their symptoms worsened over the nursing period.

Importantly, 16.9% (n=30/177) of DMER respondents stopped breastfeeding because of DMER symptoms, and a further 19.2% (n=34/177) had considered doing so.

The most frequently reported factor worsening DMER symptoms was stress which was selected by 62.1% (n=113/182), closely followed by sleep deprivation (60.4%, n=110/182). Loneliness and conflict with a significant other were also frequently cited DMER aggravation factors (49.5%, n=90/182 and 48.9%, n=89/182, respectively).

The factors most likely to ease DMER symptoms were “support from partner” and “sleep” (34.6%, n=63/182 and 29.7%, n=54/182, respectively).

**Conclusion(s):** DMER is a relatively common postpartum condition, affecting approximately one in seven lactating mothers in our study. Those with preexisting mental health and mood disorders were at elevated risk. One in six mothers with DMER stopped breastfeeding because of their symptoms. Further research and effective awareness campaigns targeting both expectant parents and their healthcare providers are needed to address this widespread but understudied problem.

**Tweetable statement:** Among 1469 surveyed new mothers, 14% experienced negative emotions with milk let-down, a ‘Dysphoric Milk Ejection Reflex’. Of those, 17% stopped nursing as a result. Baby blues, PPD, or a prior mental health diagnosis increased the likelihood of DMER.

**AJOG at a Glance:** *Why was this study conducted?:* Little is known about how many lactating parents are affected by dysphoric milk ejection reflex (DMER), predisposing factors, and what circumstances aggravate or alleviate symptoms. Similarly, the consequences of this disorder, including its effect on breastfeeding rates, are poorly understood.

*What are the key findings?:* One in seven survey respondents in our study experienced DMER. Importantly, 16.9% of those with DMER reported nursing cessation due to DMER symptoms. Those with baby blues or postpartum depression (PPD) were at significantly elevated risk for DMER, as were those with pre-pregnancy mental health conditions. 79.3% of survey respondents who had experienced DMER reported their symptoms had stabilized, improved, or disappeared prior to weaning. Sleep and partner support were the most frequently reported mitigating factors.

*What does this study add to what is already known?:* This study is the first to identify risk factors for DMER and explores DMER duration, persistence, timing, and modifying factors in the largest cohort examined to date.

## Introduction

Dysphoric Milk Ejection Reflex (DMER) is a widespread, but understudied disorder in lactating individuals, categorized by an abrupt and often overwhelming wave of negative emotions in conjunction with a milk-letdown^123^. Affected people typically describe sudden anxiety, anger or sadness, often directly prior to milk ejection, and subsiding within 1-5 min thereafter^3^. Dysphoria is typically absent in intervals between episodes, but DMER can occur at every letdown, up to 3-4 times per feeding or pumping session^4^. DMER is clearly distinct from other mental health conditions of the perinatal and lactation period, given its paroxysmal nature and characteristic temporal association with the milk ejection reflex^3^.

While some parents and perinatal health care providers have long been aware of this phenomenon, it was first formally identified by a lactation specialist, Alicia Heise, who created a support website in 2007 (https://d-mer.org/resources). Thus far, scientific data on prevalence and associated factors of DMER is scarce. The first case report on DMER was published in 2010, followed by a few case series, two reviews, and few small studies, with less than 50 DMER-affected individuals per study^5167^.

Despite a known impact on breastfeeding decisions, large data sets on prevalence and parental experiences with DMER are lacking. To address this, and to determine risk factors and measures that alleviate or worsen DMER symptoms, we conducted a large online survey and found that women with preexisting psychiatric conditions, postpartum depression (PPD), baby blues, and a migration background are at elevated risk of developing DMER. We identify partner support and sleep as important factors mitigating DMER severity.

## Materials and Methods

We conducted a cross-sectional study using an online survey (SoSci Survey GmbH, Munich, Germany). Inclusion criteria for participation were (1) having a child under the age of 18 months and (2) having nursed that child for any length of time. The survey period was June to September 2023. The link, as well as a flyer with a QR code, were widely distributed across Germany in places frequented by parents of young children, including community centers, playgrounds, “mommy-and-me” classes, postpartum exercise classes, and the offices of pediatricians, obstetricians, midwives, and pelvic floor physiotherapists. An online link to the survey was also shared in parenting-oriented online spaces.

Survey respondents were first asked for demographic data, medical and obstetric history, and experience with baby blues or PPD.

At the end of this section of the survey, participants were shown a slide introducing DMER and describing typical presentation (Suppl. 1). Survey participants were then asked if they had experienced these types of symptoms. Those who indicated “yes” were asked to fill out a second portion of the survey interrogating the specific symptoms they experienced, their duration over time, as well as mitigating and aggravating circumstances. Thus, our survey structure did not pre-select for DMER-interested or aware participants, as recruitment was conducted without mentioning the specific survey topic.

Data were analyzed in the open-source software R with the following packages: tidyverse, ggplot2, ggally, and Imtest^8^. We performed a risk factor analysis using logistic regression. The outcome of interest was the probability of developing DMER. The risk analysis required some method of feature selection, as the numbers of predictors was initially well over 20. We used the Akaike Information Criterion (AIC) to determine which variables significantly contributed to the model. We checked the AIC of the model after removing each predictor individually, and permanently removed the one that most decreased the AIC. We repeated this until the AIC no longer decreased by removing a variable, and performed a likelihood ratio test with the original model to make sure the final model with less features was comparable to the initial one. To determine if a predictor significantly affected the outcome, we fixed a significance level of 0.05.

For all survey data, we report on percentage of respondents who answered the individual question. Thus, total number of respondents for each question may vary, and this is noted in the respective figure legends.

This study received approval from Hannover Medical School Ethics Committee (10702_BO_K_2023) on May 25^th^, 2023.

## Results

Of the n=1469 respondents who completed the survey, n=209 (14.2%) reported experiencing DMER. The demographic details of this cohort are described in Table 1. The mean age was 31 with a range of 18-48 years. 100% identified as female, so we will refer to them as “women” and “mothers” in our text. The delivery mode was vaginal in 63.3% (n=873/1380), operative vaginal in 10.6% (n=146/1380), and cesarean in 26.2% (n=361/1380) for the most recent birth. Most respondents, 63.7% (871/1390) had one child. 64.0% (n=937/1464) were married, and 30.2% (n=442/1464) reported living in long-term partnerships, while 5.1% (n=74/1464) were single. The majority (n=1123/1468, 77.5%) had a degree of higher education, such as a bachelor degree or a qualification from the German apprenticeship system (Table 1). The income distribution in our survey cohort largely reflected that of the general population^9^, with the largest subgroup within the cohort (28.5%. n=405/1421) earning 40,000-59,000 € per year per household after taxes (Table 1).

**Table 1:** Respondent Characteristics (n=1469) No higher education includes respondents whose highest educational level is a high school diploma (in Germany: Abitur) as well as those with diplomas obtained after less than 12 years of schooling (Hauptschulabschluss, Mittlerer Schulabschluss) as well as those who left school without a diploma. Degree of higher education includes respondents with a degree higher than a high school diploma (in Germany: Abgeschlossene Ausbildung, Bachelor or Master, Diplom oder Promotion).

| Characteristics | Count | % |
| --- | --- | --- |
| **Sex** |  |  |
| Female | 1469 | 100% |
| Male | 0 | 0% |
| **Marital Status** |  |  |
| Married | 937 | 64% |
| Single | 74 | 5% |
| Long-term partnership | 442 | 30% |
| Divorced | 10 | <1% |
| Widowed | 1 | <1% |
| **Number of children** |  |  |
| 1 | 871 | 63% |
| 2 | 417 | 30% |
| 3 | 82 | 6% |
| 4+ | 20 | 1% |
| **Education level\*** |  |  |
| No higher education | 345 | 23% |
| Degree of higher education | 1123 | 77% |
| **Net household income in Euro** |  |  |
| <20,000 | 86 | 6% |
| 20,000-39,999 | 265 | 19% |
| 40,000-59,999 | 405 | 29% |
| 60,000-79,999 | 356 | 25% |
| 80,000-99,999 | 161 | 11% |
| 100,000-119,999 | 72 | 5% |
| 120,000-139,999 | 31 | 2% |
| 140,000> | 45 | 3% |
| **Immigration background** |  |  |
| Identify as having immigration background | 150 | 10% |
| Do not identify as having immigration background | 1315 | 90% |
| **Partner parental leave** |  |  |
| Yes | 743 | 51% |
| No | 722 | 49% |
| **Mode of birth** |  |  |
| C-section | 361 | 26% |
| Vaginal | 873 | 63% |
| Operative vaginal | 146 | 11% |
| Characteristics | Median | IQR |
| **Current age of respondent in years** | 31 | 6 |
| **Age at birth of first child in years** | 29 | 7 |
| **Current age of youngest child in months** | 10.5 | 9 |
| **BMI** | 24.46 | 6.54 |

It is important to note that in Germany, having a “migration background” reflects an identity rather than a specifically defined ethnic population. People who identify as having such a background can be recent arrivals or German-born citizens who have familial and historic ties to immigrant communities. Respondents to our survey who self-identified in this group had a higher risk of developing DMER in our logistic regression analysis (Figure 1).

**Figure 1:**
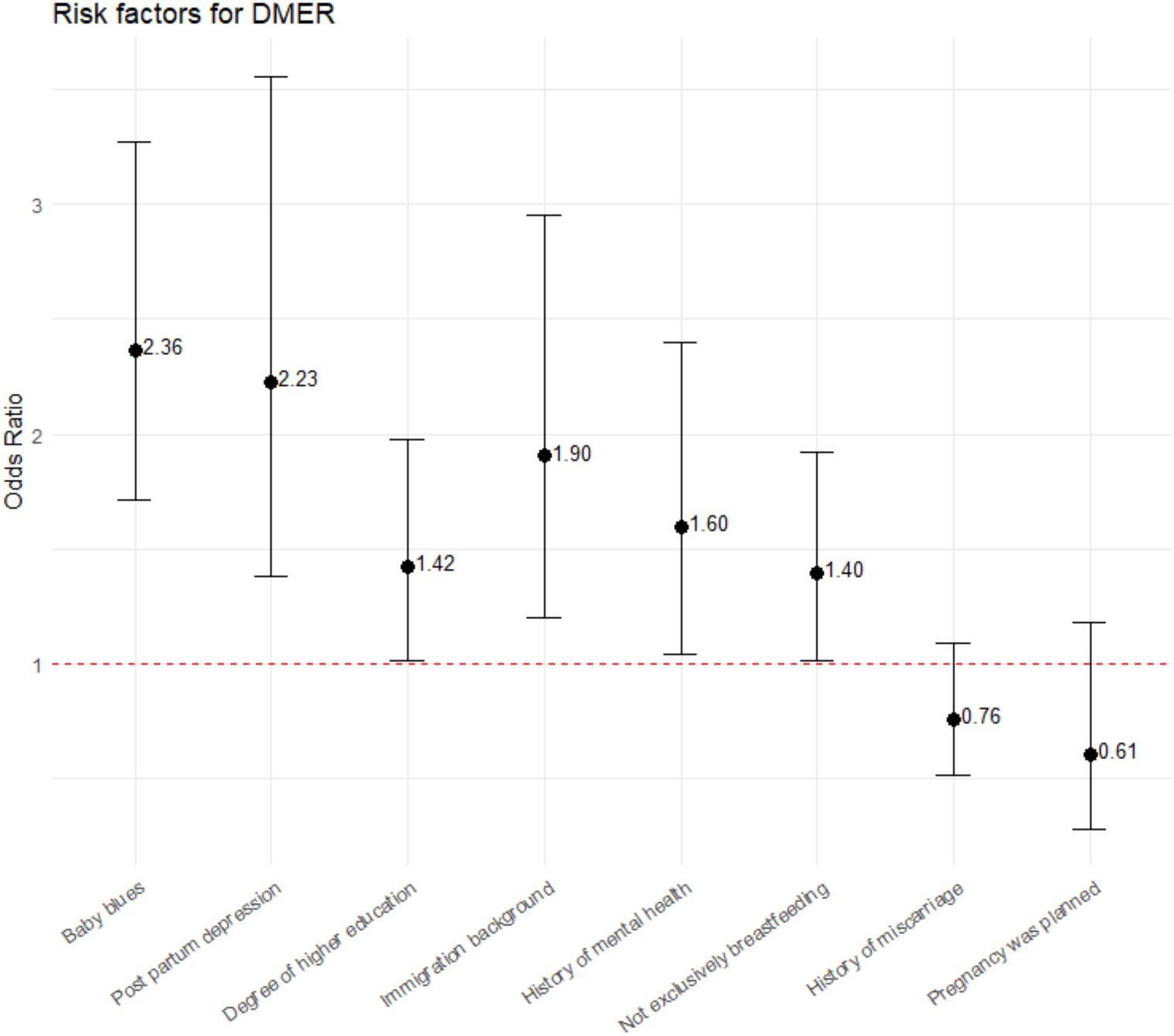
Risk factors for DMER. Odds ratios for the variables in the logistic regression and their 95% confidence intervals. An odds ratio whose confidence interval does not include 1 is considered to be significantly elevated for that variable.

Analyzing potential risk factors across the DMER and non-DMER groups, we found a high co-incidence of DMER with both PPD and baby blues (Fig 1). In each case, people who had either of these conditions were more than twice as likely to report DMER as those who did not. Similarly, mothers who reported having been diagnosed with a mental health disorder prior to pregnancy carried an elevated risk for DMER, while those with a previous history of a somatic illness did not. We identified a higher risk of DMER in those with a post-secondary degree. DMER was also more frequent in mothers supplementing their nursing with formula, likely reflecting the fact that DMER during breastfeeding led to alternative feeding choices in these families.

Marital status, household income, maternal age at birth of first child, BMI, history of medical illness, mode of birth, and sex of infant did not affect likelihood of developing DMER. Similarly, there was no change in DMER risk related to whether a pregnancy was planned or not, nor whether it was the result of assisted reproductive technology. Also statistically insignificant were a history of pregnancy or birth complications, including prematurity, a history of induced or spontaneous abortion, medication use during pregnancy, and hospitalization of the infant or birthing parent. Overall, 50.7% of non-birthing partners took parental leave for at least the first four weeks after birth, but we found no significant effect of partner parental leave on the development of DMER.

Among the n=209 women who reported experiencing DMER, we sought to explore and better define the parameters of this condition. 57.7% (n=113/196) reported their symptoms lasted for 1-5 minutes, with 18.9% (n=37/196) stating shorter duration than one minute and 23.5% (n=46/196) reporting durations above five minutes per episode (Fig 2).

**Figure 2:**
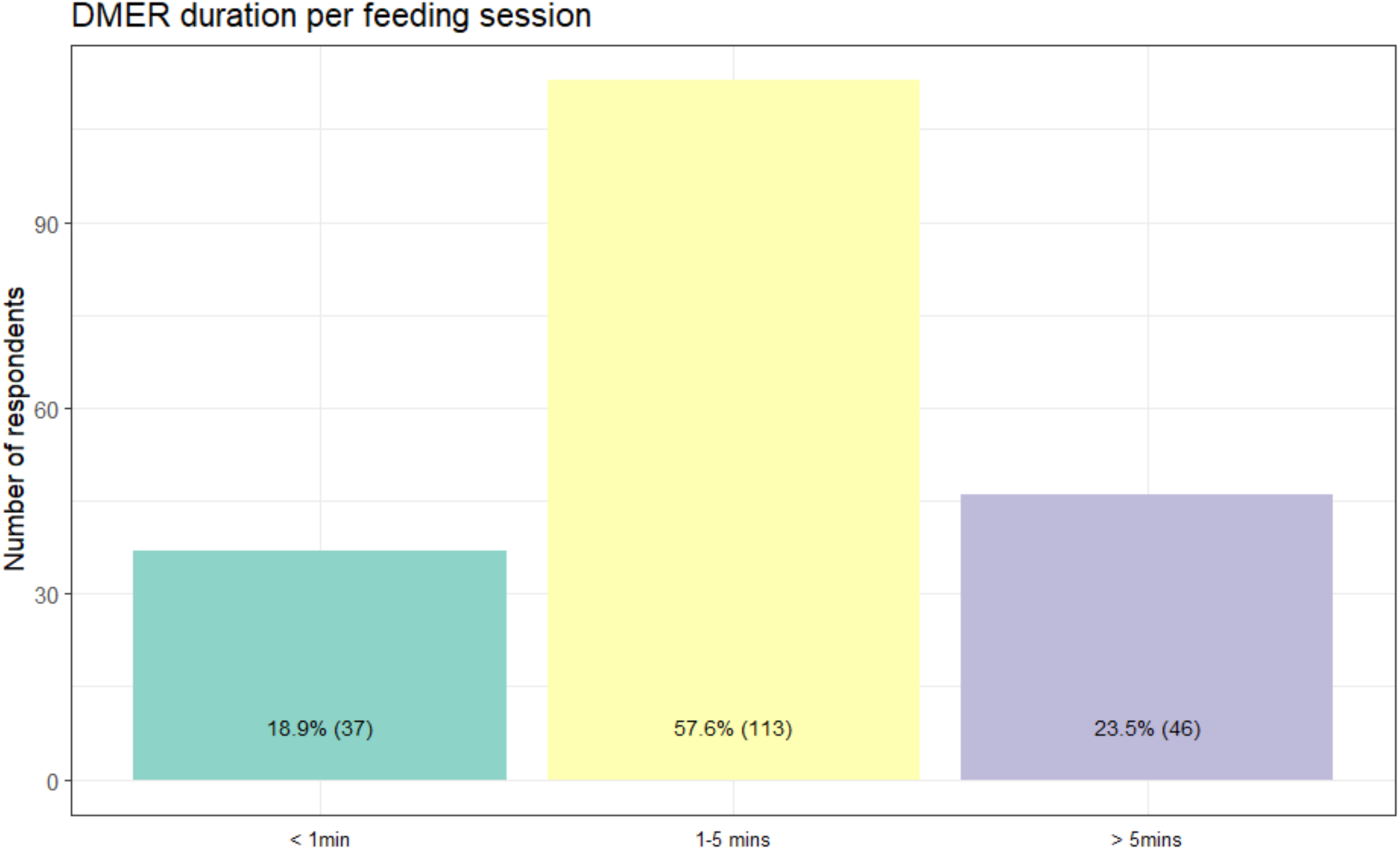
DMER duration per feeding session. Duration of the symptoms of DMER in the women surveyed that did suffer from DMER (n=196).

When asked to select feelings experienced during DMER episodes from a list of twenty-four emotions, most respondents choose between one and three. The most common were, in descending order: tense, hypersensitive, frustrated, irritable, overwhelmed, sad, and lonely (Fig 3). Similar emotions were more likely to be selected together, e.g. a high level of overlap between “angry” and “annoyed”, or “irritable” and “frustrated” occurred.

**Figure 3:**
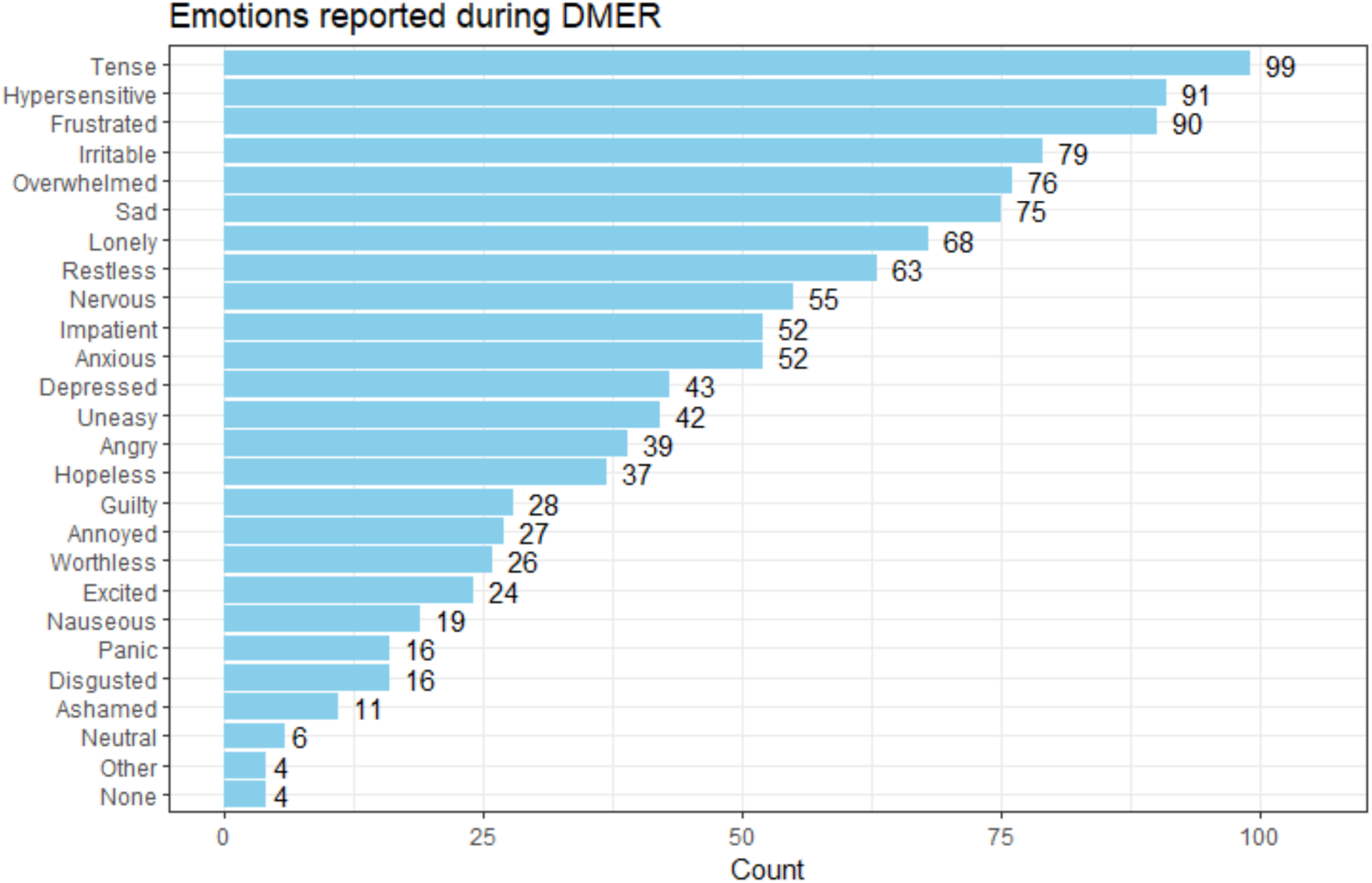
Emotions reported during DMER. Number of times each emotion was selected by the 209 respondents that experienced DMER. More than one emotion could be selected by each respondent.

85.9% (n=158/184) of mothers in the DMER group used a pump to express breastmilk (Fig 4). Of these, 44.3% (n=70/158) experienced milder DMER symptoms while pumping compared to nursing, and in 12.7% (20/158), pumping caused no DMER symptoms whatsoever. In only 5.7% (n=9/158) pumping caused more severe symptoms than nursing, and 12.0% (n=19/158) had symptoms only when pumping. For 25.3% (n=40/158) symptom intensity was equal during pumping and nursing.

**Figure 4:**
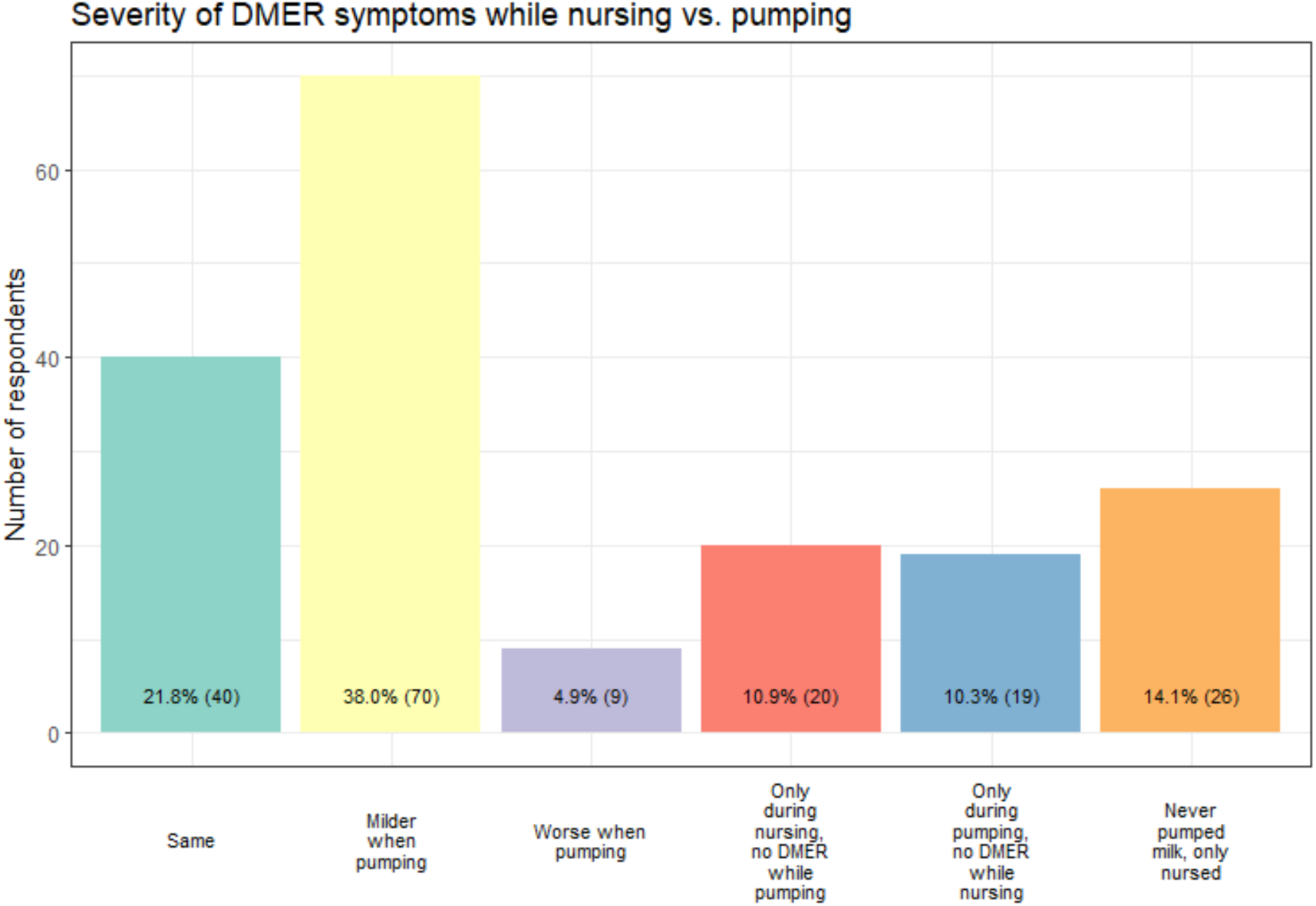
Severity of DMER symptoms while nursing vs. pumping. Comparison of nursing and pumping among women that experienced DMER (n=184 respondents answered question, n=158 used a breast pump, allowing comparison pumping vs nursing).

Regarding the timing of symptom onset, 73.3% (n=132/180) of respondents reported experiencing DMER only at the beginning of a nursing session (Fig. 5). Another 11.1% (n=20/180) experienced symptoms during every letdown while nursing. Less frequently, mothers reported DMER during initiation of pumping only (n=8/180, 4.4%), or during every letdown in a pumping session (n=2/180, 1.1%). 10.0% of women (n=18/180) reported symptoms during every letdown, regardless of whether they were pumping or nursing.

**Figure 5:**
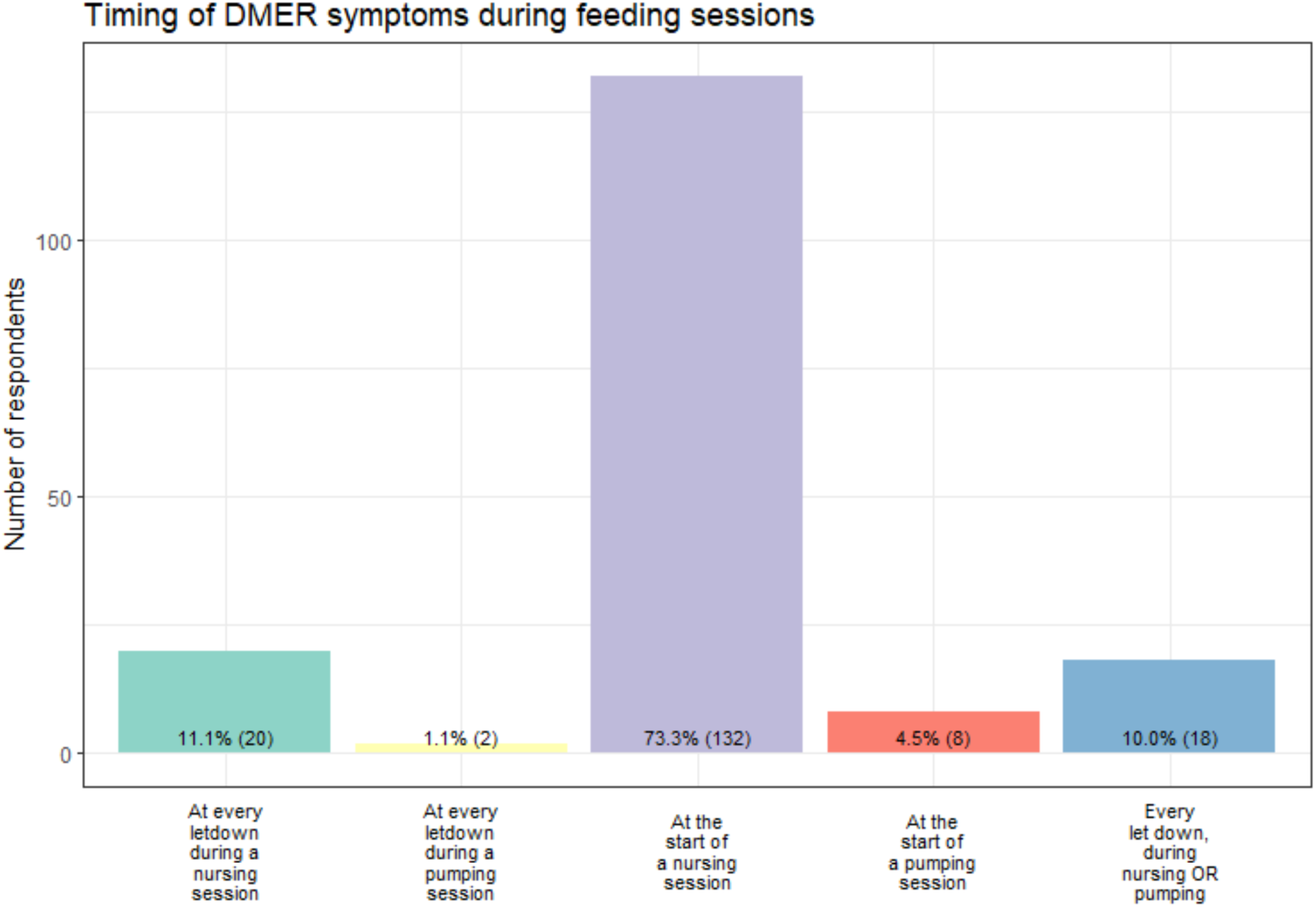
Timing of DMER symptoms during feeding sessions. Experience of timing of DMER during nursing and pumping sessions (n=180).

Regarding symptom persistence and severity between birth and breastmilk weaning, almost half of respondents in the DMER group, 40.2% (n=72/179), reported that their symptoms remained stable, in 29.6% (n=53/179) symptoms decreased, and in 9.5% (n=17/179) they disappeared completely. In contrast, 17.9% (n=32/179) of mothers reported that DMER symptoms became worse over the course of the lactation period (Fig 6).

**Figure 6:**
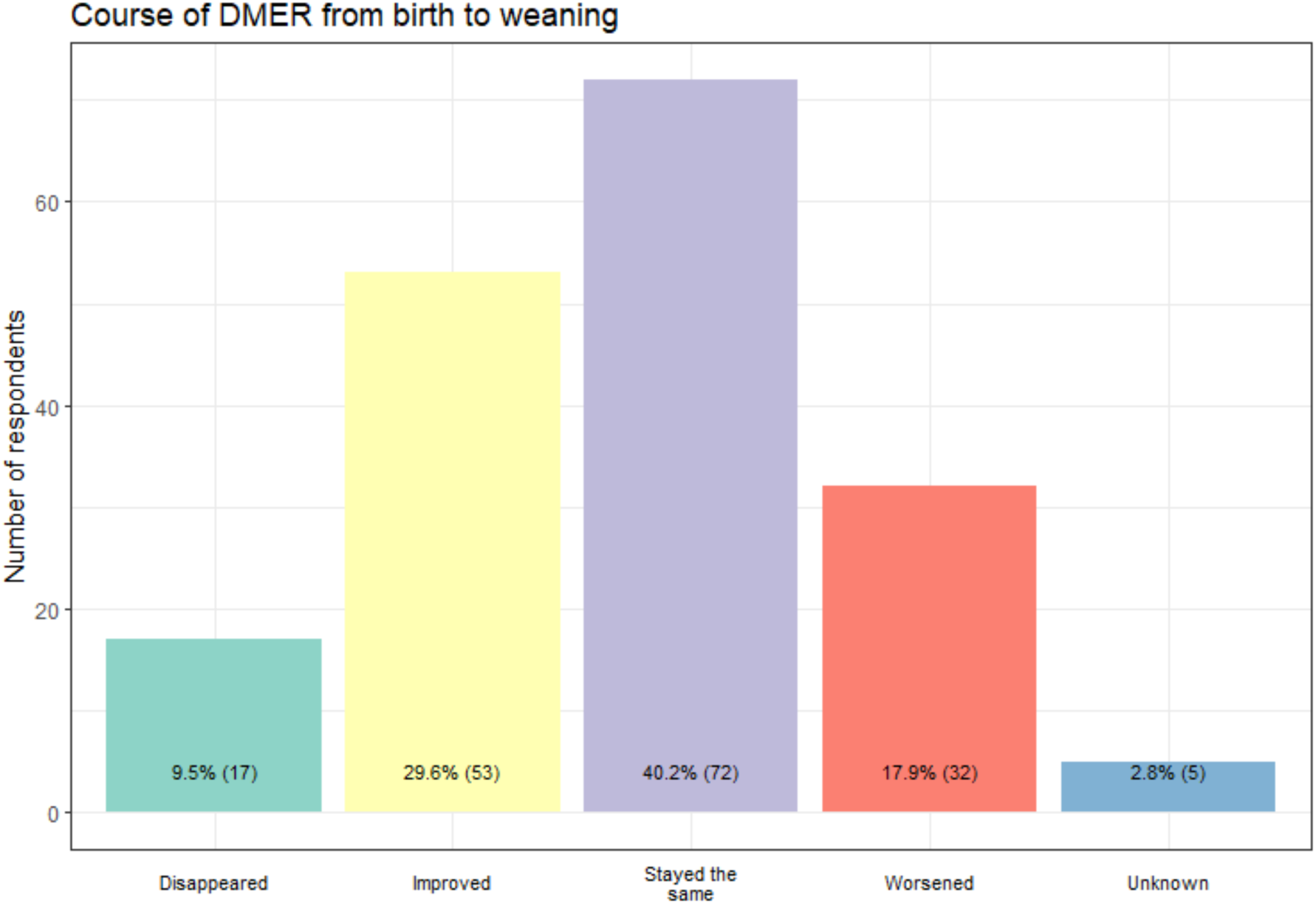
Course of DMER from birth to weaning. DMER symptom course over the period from birth to cessation of breastfeeding/pumping (n=179).

Importantly, 16.9% (n=30/177) of DMER respondents stopped breastfeeding because of their DMER symptoms, and a further 19.2% (n=34/177) had considered doing so. Additionally, 2.8% (5/177) stopped nursing a previous child due to DMER (Fig 7).

**Figure 7:**
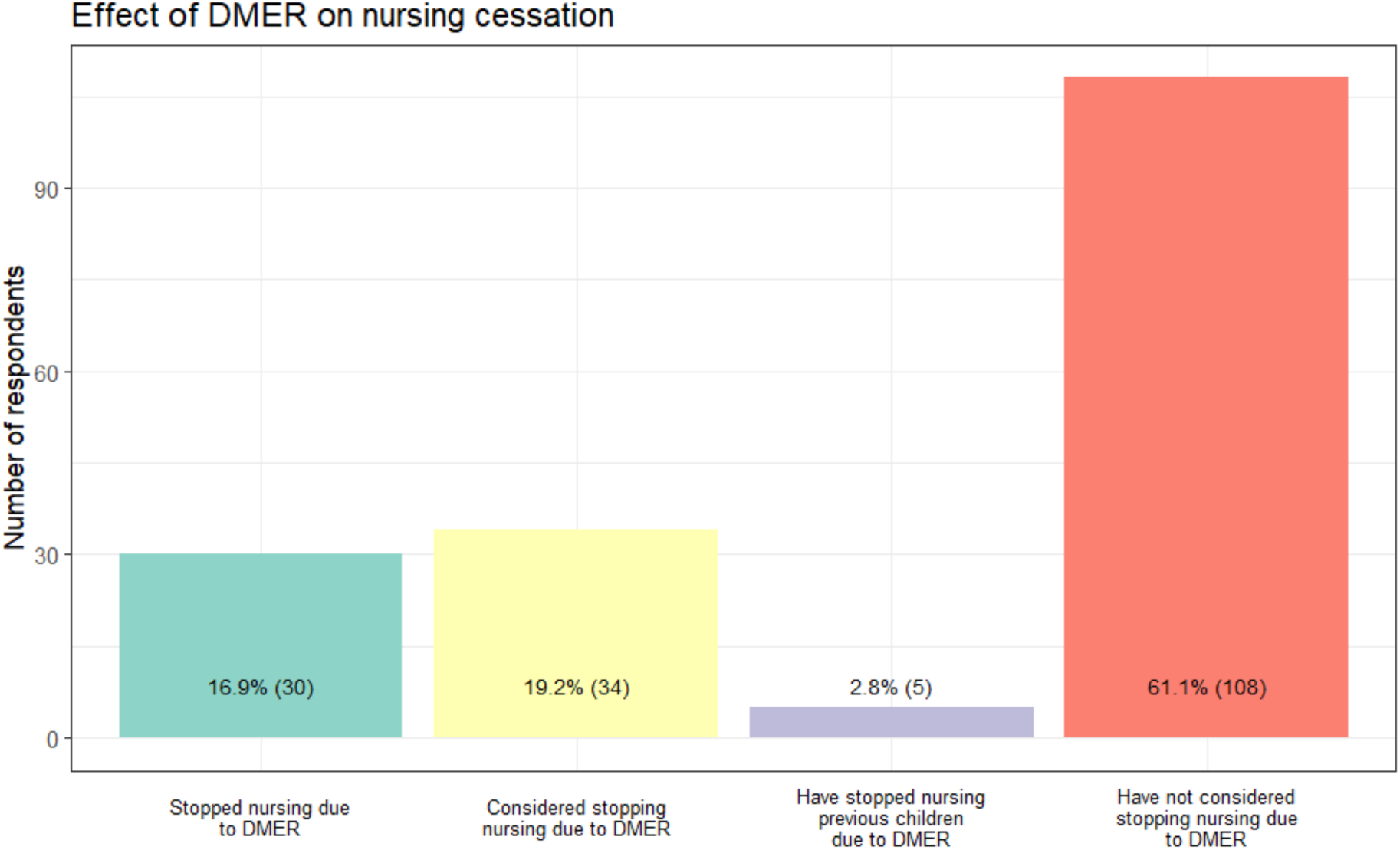
Effect on nursing cessation. Effect of DMER on infant feeding choices (n=177).

When asked which factors worsened or exacerbated DMER symptoms from a list of 10 items (suppl. Fig. 8), the respondents selected a mean of 2.7 answers. The most frequently chosen were stress (n=113/182, 62.1%), closely followed by sleep deprivation (n=110/182, 60.4%, Fig. 8). Loneliness and conflict with a significant other were also frequently reported (n=90/182, 49.5% and n=89/182, 48.9%, respectively). Interestingly, a relevant proportion (n=47/182, 25.8%) reported that shorter periods between feedings aggravated DMER symptoms.

**Figure 8:**
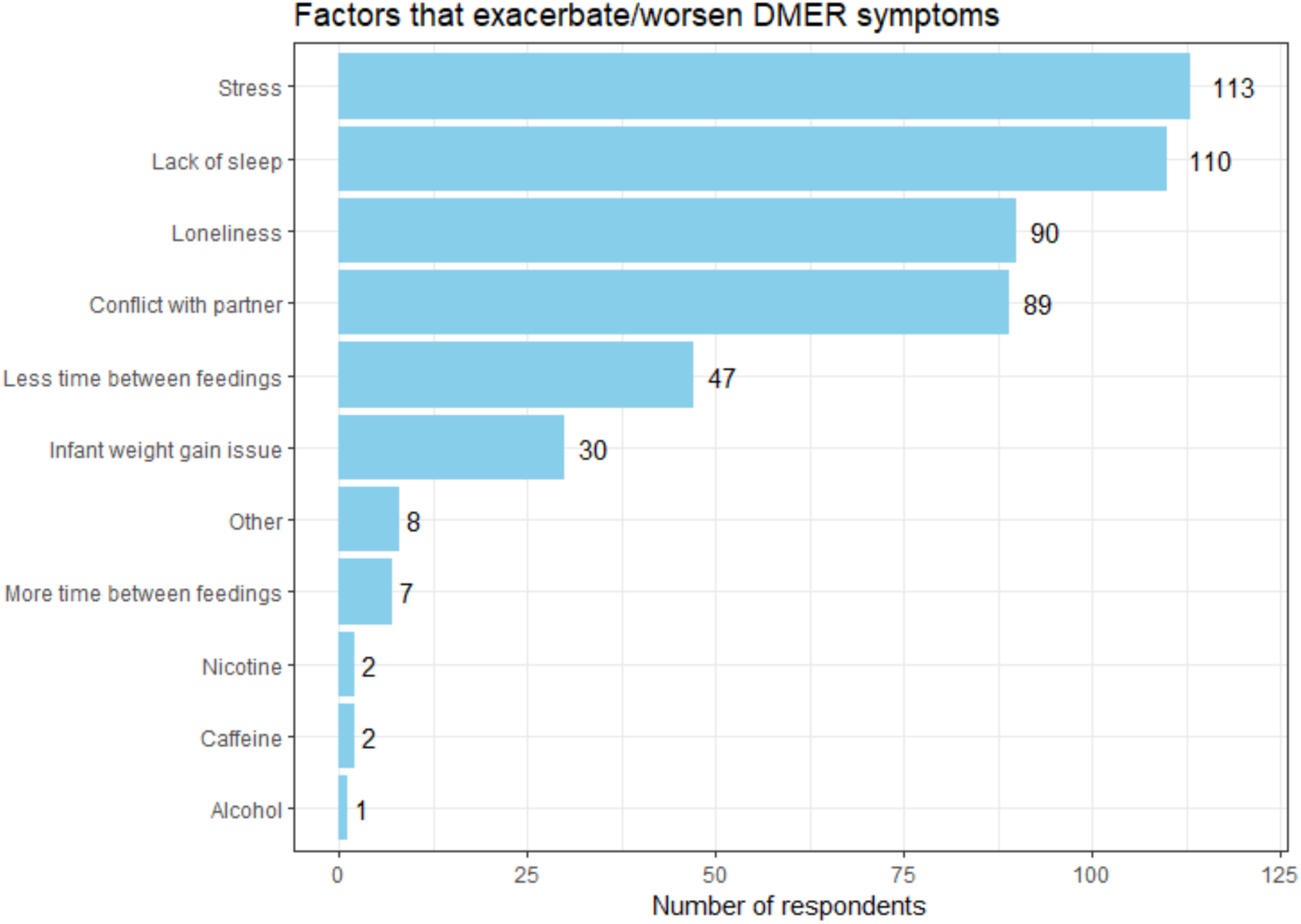
Factors that exacerbate/worsen DMER symptoms. Factors reported by women experiencing DMER that worsened symptoms. More than one answer could be selected (n=182).

Next, we asked mothers affected by DMER about factors that alleviated DMER symptoms (18 answer options, Fig. 9). Here, the mean number of selected mitigating factors was 2.3, with “support from partner” and “sleep” most frequently selected as easing factors (n=63/182, 34.6% and n=54/182, 9.7%, respectively, Fig. 9).

**Figure 9:**
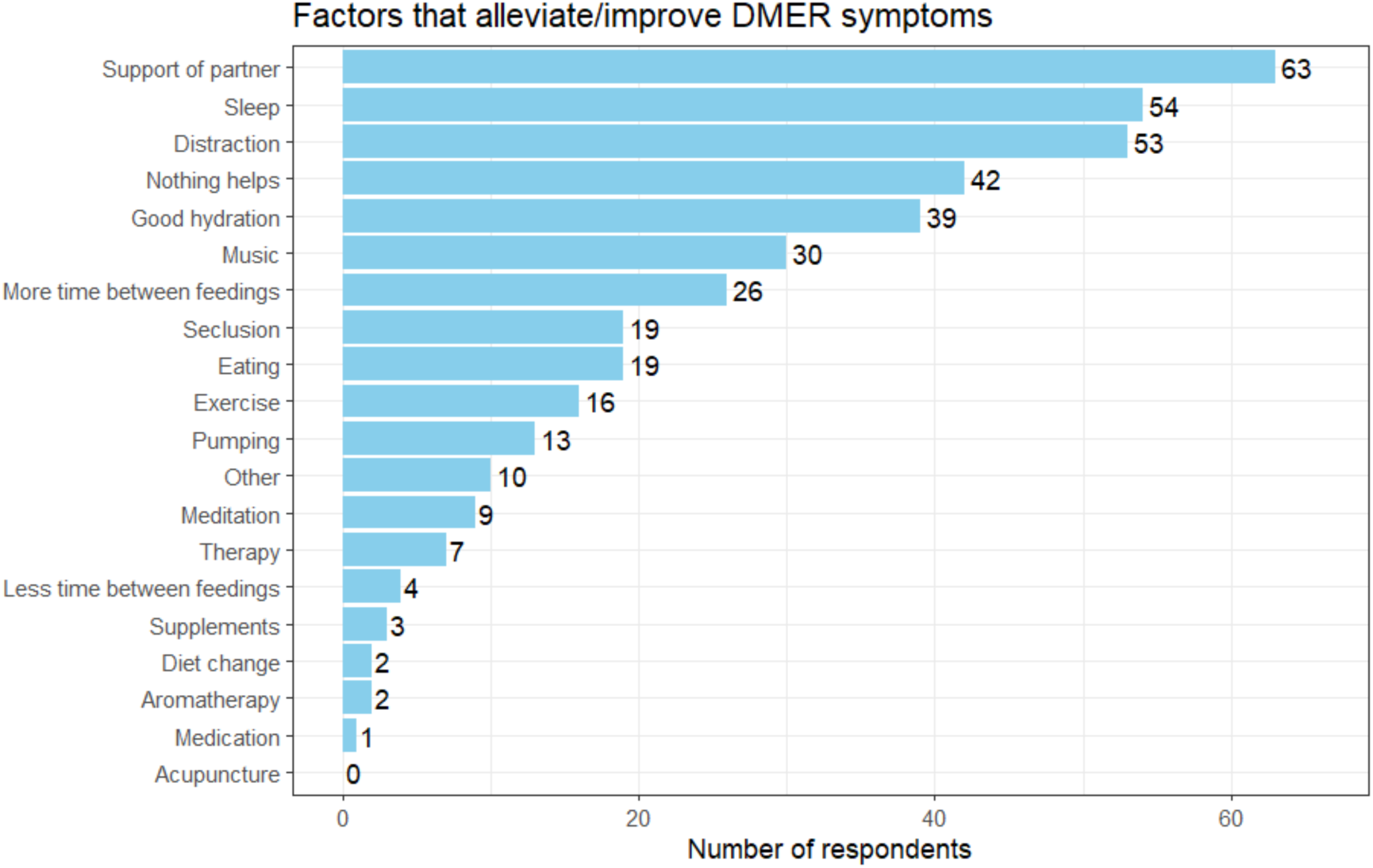
Factors that alleviate/improve DMER symptoms. Factors reported by women experiencing DMER that improved symptoms. More than one answer could be selected (n=182).

Distraction, drinking fluids, music, and a longer time between feedings were also reported to improve DMER (n=53/182, 29.1%; n=39/182, 29.4%; n=30/182, 16.5%; n=26/182, 14.2%,respectively). Importantly, however, the fourth most frequently chosen answer was “nothing helps” (n=42/182, 23.1%).

## 4. Comment

### a. Principal Findings

In our sample, 14.2% (n=209/1469) of mothers reported having DMER. Those with either PPD or baby blues were twice as likely to experience DMER, and a history of mental health conditions prior to pregnancy also elevated the DMER risk. Higher education level and migration background also increased the likelihood of developing DMER in our cohort. Symptoms were exacerbated by stress, sleep deprivation, and conflict with a significant other. Respondents reported the most helpful factors in alleviating or mitigating DMER were sleep and support from a partner. Longer periods between feedings were also associated with improved symptoms, while more frequent feeds increased severity. While more than a third of mothers experienced reduction of DMER symptoms over the course of the lactation period, almost every fifth mother experienced a worsening of her DMER over the course of her time nursing. Importantly, one in six women stopped breastfeeding due to DMER.

### b. Results in the Context of What is Known

Because DMER is a relatively recently described phenomenon, reliable data on basic characteristics such as its prevalence and persistence over time are extremely scarce^10^. To the best of our knowledge, this is the largest study to date, and we confirm a high prevalence of DMER amongst lactating mothers using an unselected sample of breastfeeding women.

The observed rate of 14.9% is in the middle of the wide range of previously published literature. The first study, a pioneering work by Ureño et al, included a retrospective chart review of 164 women attending their 6-8 week postpartum visit, 9.1% of which reported DMER-like symptoms^3^. These authors also conducted an online survey in 99 people recruited from a DMER support website and described the demographic data, obstetric history, and DMER symptoms in this cohort. The authors found that 35% of respondents had either stopped breastfeeding due to DMER or were considering doing so. The proportion of mothers ceasing nursing or considering doing so in our study is remarkably similar (36.2%). This finding confirms that DMER is a highly relevant factor in nursing decisions.

Another smaller study from the United States, included 200 women between 4 and 12 weeks postpartum and found 12 individuals with DMER, a rate of 6%^11^. This study found an association between DMER and a higher score on the Edinburgh Postnatal Depression Scale. Interestingly, pre-existing depression or anxiety were not associated with DMER, but other mood disorders were. However, with a DMER sample size of twelve, caution must be exercised in interpreting these results. A much a higher rate of DMER occurrence, 23.3% was found in a Japanese study of 202 women^12^. This study, however, may suffer from recall bias, as it was conducted in the context of the 3-year well-child visit, increasing the time between breastfeeding initiation and survey time point. The authors describe a relationship between DMER and maternal age, with those under 35 years of age more likely to be affected. In our larger study, we did not find a correlation between DMER and maternal age, either at the time of surveying or at birth of first child. Our finding that higher educational attainment increased DMER risk was also in contrast to the Japanese cohort, where educational background did not affect likelihood of developing DMER. Divergent findings in different countries are likely explained by the complex interplay of personal and cultural factors in determining the perinatal experience and reporting.

### c. Clinical Implications

Awareness of DMER among providers of obstetric health care is improving, but the lack of robust data in this area limits evidence-based management recommendations. Based on our findings, DMER appears to affect a similar number of mothers as PPD^13^ ^14^ and significantly impacts nursing practices. As such, we believe that postpartum care should routinely include information and support for DMER alongside other postpartum mental health disorders.

Providers should be especially alert when birthing parents have a preexisting history of psychiatric disease or are experiencing PPD or baby blues. The finding that a migration background elevates the risk of developing DMER is novel and can aid clinicians in identifying a vulnerable group at particular risk.

Regarding management strategies, our data finds that a supportive partner, sleep, and distraction during nursing may alleviate symptoms. Longer periods between feedings were associated with symptom improvement, and could represent a useful approach, if infant nutritional needs allow. Around half of mothers with DMER in our cohort who used a breast pump reported milder or absent symptoms while pumping as compared to nursing, suggesting another mitigating strategy. However, pumping itself may be associated with stress and earlier breastfeeding cessation, so the value of this approach may be limited^15^.

Of special concern is the finding that one in six of DMER patients stopped nursing due to their symptoms, and a further fifth of these mothers were considering doing so. While all parents should make infant feeding decisions based on their personal preferences, this high rate of cessation among parents who initially wished to breastfeed underlines the high burden associated with DMER.

Our results add to the body of evidence that provides an essential foundation for decision making among health care providers and parents. For example, there is no previous published data assessing the likelihood of spontaneous regression of DMER, despite the obvious relevance of this question to affected parents. Our study is the first to address DMER persistence over time, and our finding that symptoms improved or disappeared in around 40% of women over their lactation period, with almost 10% experiencing full resolution of DMER symptoms may help inform the choices of affected families.

### d. Research Implications

Many epidemiological and pathophysiological questions on DMER remain unanswered. In our study, a higher level of education or migration background were associated with increased risk for DMER, but net household income and age of mother were not. More research is needed to interrogate the relationship between socioeconomic factors and DMER risk, as well as the interplay with PPD and baby blues. Additionally, while we identified partner support and sleep to be relevant factors once DMER developed, having a partner at home full time for the 4 weeks following birth was not associated with a lower risk of developing DMER in our study. This may be because parental leave by the non-birthing parent is more common in high-income households, possibly veiling an effect in our study. The role of the support systems in DMER prevention and mitigation requires further study, as do the interesting differences experienced by DMER affected mothers during nursing and pumping. Indeed, a better understanding of the pathophysiological mechanisms underlying DMER would go a long way towards developing evidence-based recommendations.

### e. Strengths and Limitations

The primary strength of our study is the large sample size of unselected respondents, with over one thousand four hundred women surveyed and over two hundred mothers with DMER included in our analysis. The DMER-neutral study recruitment strategy allowed for data collection from a large sample of parents with breastfeeding experience without preselecting for women with an interest in, or awareness of, DMER. However, given the online nature of the questionnaire, selection bias is likely, as parents willing to participate in a health questionnaire and those able to scan a QR code to conduct an online survey are likely different from those who do not. For example, mothers with migration background were underrepresented in our study as compared to the general population. Another limitation of our study may lie in the fact that symptoms were self-reported. An element of recall bias is probable, as we included mothers who had given birth within the previous 18 months. It is possible that those currently nursing a newborn had a more accurate memory of early breast-feeding details and experiences than those with older children.

### f. Conclusions

We present the largest and most comprehensive data set on DMER published thus far. DMER is widely prevalent and responsible for significant rates of nursing cessation. We recommend all birthing parents be counseled and screened for DMER alongside postpartum mental health disorders. Further research is urgently needed to establish validated clinical tools as well as to interrogate the pathophysiological mechanisms underlying this understudied disorder.

## Data Availability

All data produced in the present study are available upon reasonable request to the authors

## Acknowledgments

We gratefully acknowledge the participation of all families who participated in our study and shared their experiences with us for the purposes of research. We thank Simon Ritter for helping with questionnaire programming and Ioannis Liolios for reviewing the statistical methods section.

## Survey (Translated from the original German)

**Do you have a child under 18 months old?**

Yes No

If Yes ◊ Continue

If No ◊ Survey ends, straight to “Thank you for participating” page

**How old is your youngest child in months?**

[Please select]

**How long did you breastfeed your youngest child?**

[Please select]

**Did you carry and gave birth to the baby?**

Yes No

**How was your child fed milk?**

**If you have multiple children, please answer regarding your youngest child.**

Exclusively breastfed Exclusively bottle fed from birth Both breastfed and bottle-fed.

**What is your current marital status?**

Single

In a long-term partnership, not married

Married

Widowed Divorced

Registered civil partnership

Registered civil partnership, partner deceased

Registered civil partnership, dissolved

**What is your highest school or university degree?**

No school leaving certificate

Vocational school leaving certificate (Hauptschulabschluss, at least 9 years of schooling)

Intermediate school leaving certificate (Realschulabschluss, 10 years of schooling)

Secondary school leaving certificate, (Abitur 12-13 years of schooling, qualification for University entry)

Degree from vocational training program

Completed Bachelor’s degree or equivalent

Master’s degree

State examination (for example, Lawyers, Teachers, Pharmacists)

Diploma and Magister (for example, Engineers, Architects, Artists)

Doctorate

**Which statement applies to you?**

Multiple choice possible.

I am currently working part-time.

I am currently working full-time.

I am currently on parental leave.

I am currently on maternity leave.

I am currently studying.

I am currently in training.

I am currently unemployed.

**What was the total net income in euros of all members of your household last year?**

0 – 19.999

20.000 – 39.999

40.000 – 59.999

60.000 – 79.999

80.000 – 99.999

100.000 – 119.999

120.000 – 139.999

140.000 or more

**Do you have a migration background?**

Yes

No

**How old are you in years?**

**How tall are you in cm?**

**How much do you weigh in kg?**

**Do you have one or more known illnesses?**

Examples include high blood pressure, diabetes, autoimmune diseases, thyroid problems or cancer, etc.

Yes

No

**Do you take medication regularly?**

This includes taking vitamins, folic acid or iron.

Yes

No

**Have you been diagnosed with a mental illness by a doctor or psychotherapist?**

Yes

No

**If above question “yes” ->**

**What medication do you take regularly?**

**How many pregnancies have you had that ended in a live birth?**

One

Two

Three

Four

Five

More than five

**How old were you when your youngest child was born?**

[Please select]

**What is the sex of your youngest child?**

Female

Male

**How did you give birth to your youngest child?**

Vaginal

Assisted vaginal (e.g. forceps-assisted delivery) Caesarean section

**Did you carry multiples during your most recent pregnancy?**

Yes

No

**Was the pregnancy of your youngest child planned?**

Yes

No

**Was the pregnancy with your youngest child the result of assisted reproductive technology/fertility treatment?**

Yes, we became pregnant through fertility treatments/medical assistance.

No, we became pregnant naturally.

**Did you experience any complications during the pregnancy of your youngest child?**

Examples of possible pregnancy complications are blood clots/thrombosis, hemorrhage, hypertension, gestational diabetes, severe vomiting (Hyperemesis Gravidarum), pre-eclampsia, eclampsia, HELLP-syndrome, cervical insufficiency etc.

Yes

No

**Was the birth of your youngest child premature?**

A premature birth is a delivery before the 37th week of pregnancy. Yes

No

**Did you regularly take medication during the pregnancy of your child under 18 months of age?**

This also includes taking vitamins, folic acid or iron.

Yes

No

**Did you experience any complications during the birth of your youngest child?**

Examples of birth complications are labor disturbances, shoulder dystocia, positional anomalies, umbilical cord complications, premature detachment of the placenta, haemorrhage, malposition of the cervix, non-discharge of the placenta, abnormal CTG (baby’s heartbeat), etc.

Yes

No

**Was your partner at home continuously for the first four weeks after the birth of your youngest child, for example due to parental leave?**

Yes

No

**Did you have a prolonged hospitalization around the birth of your youngest child due to your own health?**

Yes, I was hospitalized for more than a week before the birth of my youngest child.

Yes, I was hospitalized for more than a week after the birth of my youngest child.

Yes, I was hospitalized for longer than a week both before and after the birth of my youngest child.

No, I was not in hospital for longer than planned for the birth.

**Was your youngest child hospitalized for longer than planned after the birth?**

Yes, my baby had to stay in hospital for less than a week after the birth for observation or treatment.

Yes, my baby had to stay in hospital for observation or treatment between one week and one month after birth.

Yes, my baby had to stay in hospital for observation or treatment for more than a month after birth.

No, we did not have an extended hospital stay.

**Have you experienced miscarriages in the past?**

Yes

No

**Have you experienced abortion in the past?**

Yes

No

**Do/Did you pump breast milk at any time?**

Yes

No

**Did you experience “baby blues” after the birth of your youngest child?**

Baby blues usually occurs in the first 14 days after giving birth and is characterized by mood swings and crying. Other symptoms may include irritability and a feeling of being overwhelmed. This moodiness is not continuous, but alternates with periods without sadness or similar feelings.

Yes

No

**Did you experience postpartum depression after the birth of your youngest child?**

Postpartum depression is defined as a depressive episode that begins within the first four weeks after giving birth. Symptoms can include feelings of guilt, anxiety and a feeling of numbness towards the child. These feelings are continuously present and dominate everyday life without periods of temporary alleviation.

Yes

No

We would now like to ask about phenomenon that has recently become better described in medical literature called “Dysphoric Milk Ejection Reflex” (DMER). DMER is an abrupt, often overwhelming wave of negative emotions that come only during milk-letdown. These feelings can include anger, sadness or any other intense emotion.

**Have you experienced these or similar symptoms while breastfeeding your youngest child?**

Yes

No

**Did/do the DMER symptoms occur suddenly and occur only during feeding or pumping sessions?**

Yes

No

**Shortly before or during the milk letdown, I felt/feel…**

Multiple selections possible.

Anxious

Excited

Tense

Disgusted

Depressed

Frustrated

Nervous

Neutral

Panicky

Irritable

Shameful

Guilty

Sad

Hypersensitive

Restless

Impatient

Unwell

Overwhelmed

Nauseous

Annoyed

Worthless

Angry

Other

None of the above applies to me.

**I felt/feel one or more of the above symptoms:**

*Multiple selection possible*.

Only at the beginning of each nursing session.

During every letdown of a nursing session

Only at the beginning of a pumping session

With every milk let-down during pumping sessions

With every spontaneous milk let-down that occurs while neither breastfeeding or pumping.

**How long did/does DMER last for you?**

less than 1 minute between 1 and 5 minutes more than 5 minutes

**How were/are your DMER symptoms when pumping compared to those when breastfeeding?**

My symptoms are milder when pumping than when breastfeeding.

My symptoms are the same when pumping as when breastfeeding.

My symptoms are more pronounced when I pump than when I breastfeed.

I only experience these symptoms when pumping, but not when breastfeeding.

I only experience these symptoms when breastfeeding, but not when pumping.

I do not use a breast pump.

**Which statement most accurately reflects your experience?**

*Note: This question refers to improvement or worsening in DMER symptoms before weaning occurred*.

During the time between giving birth and weaning, my DMER symptoms were the same throughout.

During the time between birth and weaning, my DMER symptoms improved.

During the time between birth and weaning, my DMER symptoms disappeared completely.

During the time between birth and weaning, my DMER symptoms worsened.

I am at a point in my breastfeeding journey where I am not yet able to comment on this.

**Have you considered stopping breastfeeding because of the DMER symptoms?**

Yes, I have already stopped breastfeeding because of DMER.

Yes, I have considered/am considering stopping breastfeeding because of DMER. No, I have not considered stopping breastfeeding because of DMER.

No, not this time, but I have stopped breastfeeding other children because of these symptoms.

**Is there anything that has a positive influence on your DMER symptoms when breastfeeding your youngest child?**

Multiple selection possible.

Distraction

Pumping instead of breastfeeding

Acupuncture

Aromatherapy

Exercise

Change of diet

Food

Fluid intake

Medication

Meditation

Music

Nutritional supplements

Psychological counselling/therapy

Sleep/rest

Support from partner or others

Shortened intervals between feeding sessions Increased time between feeding sessions Seclusion

Other

No, I haven’t found anything that helps me with this.

**Is there anything that makes your DMER symptoms worse?**

Multiple selection possible. Loneliness/being alone

Caffeine

Conflict in my relationship with my partner

Lack of weight gain of the child

Lack of sleep

Stress

Shortened intervals between feeding sessions

Longer intervals between feeding sessions Alcohol

Nicotine consumption Other

